# Antibiotic use and outcomes in chronic obstructive pulmonary disease exacerbations

**DOI:** 10.1101/2025.06.20.25330010

**Authors:** Matthew Loiars, Heather Morrison, Eric Linn, Alexis Gonzalez

## Abstract

**Purpose:** The purpose of the study was to evaluate if there is a difference in clinical worsening for patients receiving different antibiotics for COPD exacerbations.

**Methods:** This was a multicenter, retrospective cohort study. Inclusion criteria were: adults whose indication for antibiotics was a COPD exacerbation; who received azithromycin, doxycycline, levofloxacin, or a beta-lactam for more than 48 hours; who received no more than 48 hours of combination initial antibiotics; and who were discharged with certain bronchodilators. The primary endpoint was clinical worsening defined by antibiotic/steroid escalation/addition. Secondary endpoints included days of antibiotic therapy, length of stay and 30-day readmissions.

**Results:** A total of 2,390 patients were identified for review with 114 patients included and 1,054 patients excluded. Patients were screened in reverse chronological order using quota convenience sampling. Clinical worsening occurred in 29% of azithromycin patients, 23% of doxycycline patients, 19% of levofloxacin patients, and 24% of beta-lactam patients; however, a Pearson Chi-Square test (χ^2^ = 0.911, *p* = 0.8227) showed no statistically significant association between antibiotic group and likelihood of escalation. Secondary outcomes that were statistically significant included the average cumulative inpatient and outpatient prednisone milligram equivalents with beta-lactams compared to azithromycin, which was 252 milligrams higher (p = 0.0161). Days of therapy (DOT) was statistically different for azithromycin compared to all of the other antibiotics.

**Conclusion:** This study concluded that there were no associated differences between groups for clinical worsening. This study showed a trend that narrow spectrum antibiotics, such as doxycycline and azithromycin, have similar outcomes as more broad spectrum antibiotics.

## Background

According to the 2025 Global Initiative for Chronic Obstructive Lung Disease (GOLD) guidelines, chronic obstructive pulmonary disease (COPD) has a global impact of 3 million deaths annually.^1^ Antibiotics are the mainstay of treatment for moderate to severe COPD exacerbations based on clinical presentation, such as increased dyspnea, increased sputum production or purulence of sputum.^1^ When indicated, antibiotics can reduce hospital duration and treatment failure through shortening recovery time and reduced risk of early relapse.^1^ The decision on antibiotic prescribing is multifactorial; these factors may include medication allergy or intolerance, known possible adverse reactions, and local resistance patterns. These antibiotics should be either an aminopenicillin with clavulanic acid, macrolide, tetracycline, or in select patients, a quinolone.^1^

Previous studies have tried to determine which of these antibiotics is preferential over another. Baalbaki and colleagues compared azithromycin to beta-lactams for COPD exacerbations and found that the beta-lactam group posed a higher risk of treatment failure.^2^ A 2021 study examined antibiotic selection on quality of life (QOL), defined as reduced exacerbations and unwanted events, and found that prophylactic macrolides improved QOL in comparison to fluoroquinolones and tetracyclines.^3^ A multicenter retrospective study compared elderly patients that received broad versus narrow spectrum antibiotics and found no difference between the groups.^5^ Another retrospective single center study found that non-fluoroquinolones had increased clinical resolution, but fluoroquinolones had shorter length of stay and inpatient antibiotic duration while both groups maintained similar rates of *Clostridioides difficile* infection.^6^

## Purpose

There are only a few studies that evaluate success between the recommended antibiotics. The primary objective of this study was to assess the resolution of COPD exacerbations requiring hospitalization regarding different antibiotic classes prescribed.

## Materials and Methods

This was an Institutional Review Board approved, retrospective, observational cohort study conducted within a four-hospital health system between February 2019 and September 2024. Study arms were divided into four groups: beta-lactams, doxycycline, levofloxacin and azithromycin.

### Patients

Patients were included if they reached these criteria: greater than or equal to 18 years of age, indication for antibiotics was a COPD exacerbation; who received azithromycin, doxycycline, levofloxacin, or a beta-lactam for more than 48 hours; who received no more than 48 hours of combination initial antibiotics; and who were discharged on both a long-acting beta agonist (LABA) and a long-acting muscarinic antagonist (LAMA) bronchodilator. Beta-lactams were defined as ceftriaxone, cefepime, piperacillin-tazobactam, ampicillin-sulbactam, cephalexin, cefdinir, cefuroxime, amoxicillin-clavulanic acid and meropenem. Subjects were identified utilizing pharmacy surveillance software VigiLanz and our electronic medical record Sunrise Clinical Manager. Manual chart review was performed to determine exclusion or inclusion based on pre-determined criteria. The algorithm for exclusion criteria checked was concomitant antibiotics outside of selected classes/antibiotics, combination antibiotics for greater than or equal to 48 hours, duration of antibiotics, if not discharged on long acting beta agonist and long acting muscarinic antagonist (LABA/LAMA) bronchodilators, concurrent infections, direct admission to the intensive care unit (ICU), mechanical ventilation on day of admission, if the patient was discharged to hospice, if the patient is not in active COPD exacerbation, patients that expired within 24 hours of admission, pregnant or breastfeeding, and if there was missing or incomplete information in the chart.

Baseline characteristics of patients included age, sex, race, body mass index (BMI), allergies, sputum PCR and lower respiratory culture results.

### Data Collection and Measurements

The justification for the order of exclusion criteria was to easily exclude patients when the initial manual chart review started and to highlight an institutional problem, combination antibiotics for greater than 48 hours. It was determined by the research team to utilize a reverse quota convenience sampling method to guarantee some data in each group due to time constraints. Using this method, the surveillance data was divided into groups by antibiotic received, then reviewed from the most recent patients until 25 patients were included. Once there were 25 patients in each group, then manual chart review would occur in the next antibiotic group.

### Outcomes

The primary outcome of the study was clinical worsening defined by antibiotic or steroid escalation/addition. Escalation was defined by the addition of another antibiotic class, broadening of antibiotic coverage within the same class (beta-lactams), and increasing frequency or dose of equivalent steroids. Secondary endpoints included 30-day readmission rates, length of stay (LOS), and total DOT.

### Statistics

Statistical analysis was performed using JMP Pro 18 (SAS Institute Inc., Cary, NC, 1989-2025) with continuous data using student’s t-test and nominal data using Chi-Square test. Based on a previous study, it was determined that sample size of 196 patients (49 patients in each group) was needed to evaluate noninferiority regarding clinical worsening, using an α value of 0.05 and a β value of 0.2 (80% power).

## Results

A total of 2,390 patients were identified for review based on the exclusion and inclusion criteria from the surveillance report, with 1,054 excluded after chart review. The most common reason for exclusion was combination antibiotics for longer than or equal to 48 hours. Patients were screened reverse chronologically using quota convenience sampling, resulting in 114 patients for analysis. The resulting four groups had 31 patients for azithromycin, 25 patients for beta-lactams, 31 patients for doxycycline, and 27 patients for levofloxacin (Table 1).

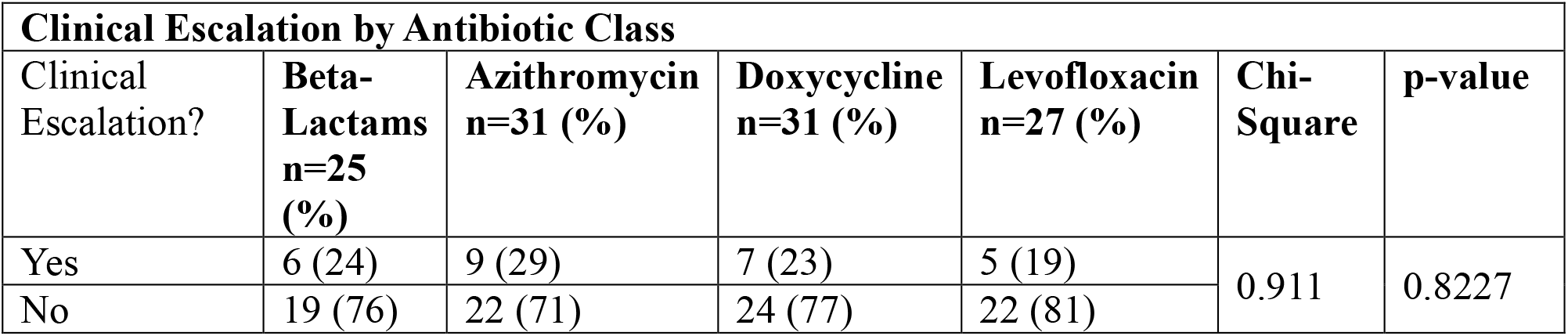
Table. 1.

Clinical worsening occurred in 9 azithromycin patients (29%), 7 doxycycline patients (23%), 5 levofloxacin patients (19%), and 6 beta-lactam patients (24%) (Table 1). The Pearson-Chi-Square value was 0.911 with a p value of 0.8227, indicating there is no statistically significant association between the antibiotic groups and the likelihood of escalation. For clinical escalation defined by antibiotic escalation, it occurred in five azithromycin patients (56%), four doxycycline patients (57%), zero levofloxacin patients (0%), and three beta-lactam patients (50%). For clinical escalation defined by steroid escalation, it occurred in six azithromycin patients (67%), four doxycycline patients (57%), five levofloxacin patients (100%), and four beta-lactam patients (67%).

Secondary outcomes that were statistically significant included the average cumulative inpatient and outpatient prednisone milligram equivalents with beta-lactams compared to azithromycin, which was 252 milligrams higher (p = 0.0161) (Table 2). DOT was statistically different for azithromycin compared to all the other antibiotics: 2.94 days shorter than beta-lactams (p < 0.0001), 2.42 days shorter than doxycycline (p = 0.0002), and 1.69 days shorter than levofloxacin (p = 0.0119).

Table 2.

**Table 2**
| **Baseline Characteristics - Difference in Prednisone Equivalents** |  |  |  |  |  |
| --- | --- | --- | --- | --- | --- |
| **Antibiotic Comparison** | **Difference** | **Std Err Dif** | **Lower CI** | **Upper CI** | **p-Value** |
| Levofloxacin vs Doxycycline | 0.92 | 101.00 | -199.24 | 201.09 | 0.9927 |
| Doxycycline vs Azithromycin | 67.33 | 97.46 | -125.81 | 260.47 | 0.4911 |
| Levofloxacin vs Azithromycin | 68.25 | 101.00 | -131.92 | 268.42 | 0.5006 |
| Beta-Lactam vs Levofloxacin | 183.84 | 106.50 | -27.22 | 394.89 | 0.0871 |
| Beta-Lactam vs Doxycycline | 184.76 | 103.14 | -19.64 | 389.16 | 0.0760 |
| Beta-Lactam vs Azithromycin | 252.09 | 103.14 | 47.69 | 456.49 | 0.0161 |
| Std = standard, Err = error, Dif = difference, CI = Confidence interval |  |  |  |  |  |

## Discussion

This study was conducted to assess the resolution of COPD exacerbations requiring hospitalization regarding different antibiotic classes prescribed. The results of the study were unable to show a statistically significant difference for clinical worsening defined as antibiotic or steroid escalation/addition. The only statistically significant outcomes for secondary endpoints showed shorter DOT for azithromycin and decreased steroids for the azithromycin group compared to the beta-lactam group. Several years prior our institution, implemented a protocol for pharmacists to get notified when patients receive 1,500 mg but still have an active order for azithromycin. It would exclude patients that are receiving azithromycin for mycobacterial infections but include patients being treated for COPD exacerbation and community acquired pneumonia. This was installed so that clinical pharmacists can intervene and discontinue the order per protocol without the patient receiving more azithromycin than needed. Another statistically significant finding was the difference in average cumulative inpatient and outpatient prednisone milligram equivalents between beta-lactams and azithromycin. There may have been a difference in this finding due to the inherent anti-inflammatory effects of azithromycin, thus lowering the need for steroids to reduce inflammation. Another reason for this finding could be selection bias from the providers choosing the patients that present worse getting a beta-lactam, thus explaining why that group would receive more steroids. While the primary outcome was not statistically significant, this study contributes real-world data to the limited evidence comparing antibiotic classes in hospitalized COPD exacerbations.

This study displayed there was no associated differences between groups for clinical worsening. While the study did not reach the power threshold in each group (49 per group), this study may show a trend that narrow spectrum antibiotics, such as doxycycline and azithromycin, have similar outcomes as more broad spectrum antibiotics.

The strengths of this study included an extensive exclusion criterion to reflect the true impact of antibiotic selection and its effect on clinical worsening. This study also included a broad definition of beta-lactams instead of just amoxicillin-clavulanic acid as per the guidelines, as real world practice does not always reflect guideline directed medical therapy. This study had several limitations. First, only CMS readmissions could be screened, which limits the visibility of the true number of patients that were readmitted for COPD exacerbation. Another limitation is the use of quota convenience sampling; this method could restrict the number of patients included and introduce selection bias into the study. This approach also required more resources to manually chart review to verify if patients were eligible to be included. The study did not account for all variables that could impact these outcomes including severity of COPD, patient adherence to prescribed outpatient antibiotics/steroids, and culture results. These limitations could impact patient outcomes significantly through patient functional status, provider antibiotic selection, and patient adherence to a once daily antibiotic versus four times daily antibiotic. Limitations were mitigated by ensuring that included patients have guideline directed medical therapy in regard to bronchodilator therapy post hospitalization and reducing concomitant infections/antibiotics.

For future studies, to address the shortcomings of this study previously mentioned, would be to include a subgroup analysis of those with different severities of COPD. This would help determine if baseline COPD disease progression played a factor in antibiotic prescribing patterns or “escalation” of antibiotics. Another improvement could be to include a subgroup analysis of the beta-lactam group to determine if there are any significant differences between specific antibiotics.

While this study did not demonstrate a difference in the effectiveness of various antibiotics in COPD exacerbation management, it did identify a difference in DOT between azithromycin and all of the other antibiotic groups as well as steroids received between azithromycin and beta-lactam groups. Further research is needed to assess antibiotic effectiveness while also accounting for COPD severity and other variables as previously listed in the limitation section. This study demonstrated that provider antibiotic selection could be improved upon and streamlined to not have combination antibiotics when pneumonia is not suspected. To further improve upon this study, sputum culture results and allergies should be taken into account to justify prescribing patterns or identify if antibiotic change is truly escalation or streamlining antibiotics.

## Data Availability

All data produced in the present study are available upon reasonable request to the authors

